# Temperature-related climate change impacts on neurodegenerative diseases: Systematic review

**DOI:** 10.1101/2025.06.02.25328777

**Authors:** Lucas Longo Ferreira, Samuel Santos Souza, Breno Medeiros, Mariana Lira Schlodtmann d’Ávila, João Pedro Lopes Pinto Loja, Henrique Maurício da Silveira, Leonardo Oliveira Nascimento, João Pedro de Godoi Moura, João Marcelo Christo Soares, Alair Augusto Sarmet Moreira Damas dos Santos

**Affiliations:** Department of Radiology, School of Medical Sciences, Federal Fluminense University

## Abstract

Climate Change poses one of the main challenges for humanity, whose impacts are wide and might be felt in areas such as human health. In this portrait, temperature implications on neurodegenerative conditions - such as Parkinson’s Disease, Alzheimer’s Disease and Motor Neuron Diseases - must be better understood. Thus, the present Systematic Review was performed by accessing the databases PubMed Central (PMC), Medical Literature Analysis and Retrieval System Online (Medline), Cochrane Library, Latin American and Caribbean Literature in Health Sciences (Lilacs), Scopus, and Embase. From the 325 papers initially identified in the field, 8 were finally selected, with the risk of bias analyzed with Cochrane’s ROBINS-E-tool. The conclusions, even though with different focus adopted by the original works, pointed that extreme heat events might worsen clinical outcomes in neurodegenerative diseases by mechanisms such as oxidative stress, neuroinflammation and, in the case of cold, changes in the phosphorylation of the Tau protein, also signaling the need for novel studies to further evaluate these findings and the public health urgency for dealing with climate change.

## Introduction

Neurodegenerative disorders are progressive degenerative conditions characterized by the functional deterioration and loss of selectively vulnerable neurons.^1^ They are incurable and debilitating diseases that have an impact on the cognitive function, affecting behavior, memory, language, learning and emotion capacity^2,3^. Neurodegenerative disorders are also responsible for shortening the life span of those affected by it^3^. Most neurodegenerative diseases have ageing as their primary risk factor, because of these nine biological hallmarks of ageing: genomic instability, telomere attrition, epigenetic alterations, loss of proteostasis, mitochondrial dysfunction, cellular senescence, deregulated nutrient sensing, stem cell exhaustion and altered intercellular communication.^4^ In addition, the identification of mutation in genes has provided important evidence for understanding the pathogenesis associated with neurodegenerative disorders. Various behaviors and environmental risk factors have been proposed for each disorder, but most associations made lack evidence. ^5^

Global warming, caused by human activities, consists on the increase in concentration of greenhouse gases, such as CO2 CH4 N2O and water vapor in the atmosphere, leading to the increase in the average surface temperature of the earth.^6^ In the past 30 years surface temperature has increased around 0.2°C per decade^7^, and in 1995 the Intergovernmental Panel on Climate Change - a multi-disciplinary scientific body established by the United Nations - concluded that the on balance an anthropogenic influence upon the global climate was now “discernible”. The expected rate of climate change over the coming century is predicted to be far greater than any natural change in the world climate since the advent of agriculture 10 000 years ago; meaning that for the first time the global impact of humankind exceeds the physical and ecological limit of the biosphere.^8^ Furthermore, it is predicted that the negative health impacts of climate change would disproportionately affect warmer and poorer regions of the world.^9^

It is known that the increased frequency and intensity of heat waves caused by global warming, have been linked to several health problems. Although the increase in prevalence of neurodegenerative diseases is well documented by literature reports, the link between global warming and the enhanced prevalence of such diseases remains elusive.^10^ Therefore, to better understand this possible correlation, it is essential to do more research on the subject. High temperatures can exacerbate issues in thermoregulation, which is already compromised in patients with neurodegenerative disorders. This can interfere with processes like excitotoxicity, oxidative stress, and neuroinflammation, all of which are related to neurodegeneration.^10^ On the other side, hypothermia can lead to increased levels of phosphorylated Tau, a protein associated with Alzheimer’s and other neurodegenerative diseases. This suggests that cold temperatures might trigger or exacerbate neurodegenerative processes by affecting Tau phosphorylation.^11^

In this sense, it urges the need for a deeper understanding on potential correlations between these, leading to the present systematic review with meta analisis. Thus, it’ll be possible to clarify and measure associations, guiding public health policies in the sense of reducing main risk factors for these conditions.

## Methodology

It was performed a systematic review on temperature-related climate change impacts on neurodegenerative diseases. This study follows the guidelines of the Preferred Reporting Items for Systematic Reviews and Meta-Analyses Protocol (PRISMA) to ensure transparency and methodological rigor, with a register on the International Prospective Register of Systematic Reviews (PRESPERO) code CRD42024504372, which protocol follows as supplementary material. The search strategy was designed to include multiple databases, such as PubMed Central (PMC), Medical Literature Analysis and Retrieval System Online (Medline), Cochrane Library, Latin American and Caribbean Literature in Health Sciences (Lilacs), Scopus, and Embase, without any search on gray literature (books, thesis, governmental relatories, etc). The temporal scope of the search includes studies published between January 1, 2019, and December 31, 2023, with a posterior complementation of studies published in 2024 using the same search strategy. Keywords were selected to cover all possible combinations between temperature-climate-related terms—such as “Climate Change,” “Global Warming,” and “Heatwaves”— and neurodegenerative diseases, including “Alzheimer’s disease,” “Parkinson’s disease,” “Vascular dementia,” “Frontotemporal lobar degeneration,” “Progressive supranuclear palsy,” “Posterior cortical atrophy,” “Dementia with Lewy Bodies,” “Amyloidosis,” “Normal Pressure Hydrocephalus,” “Creutzfeldt-Jakob disease,” “Multiple System Atrophy,” “Huntington’s disease,” “Corticobasal degeneration,” “CADASIL,” “Amyotrophic Lateral Sclerosis,” “Neurodegeneration with brain iron accumulation,” and “Neurodegeneration with brain mercury accumulation.” These combinations were arranged using the Boolean operator “AND” to refine the search results.

The eligibility criteria for inclusion in this review are as follows: randomized clinical trials, case series, cohort studies and observational studies, all of which must involve human populations. Studies focused on individuals of all age groups diagnosed with neurodegenerative diseases, particularly those specified above, and addressed exposure to environmental variables linked to climate change, such as temperature fluctuations (e.g., heatwaves and cold waves), and other relevant factors. Studies conducted on non-human populations (animal models), those that do not explore the correlation between climatic variables and neurodegenerative diseases, or those that fail to meet the specified methodologies were excluded due to lack of correlation between conclusions. The data selection process was performed by two independent blind reviewers, and the criteria for inclusion considered the availability of the full text, publication in Portuguese, Spanish, English, or French, relevance to climate change, human subjects, and a clear connection to neurodegenerative diseases. All decisions were recorded in Excel, and the CADIMA® software was used to streamline the process, being the data extraction performed using standardized forms for avoiding information bias. The statistical model - aleatory - fitted better to this review once the clinical heterogeneity is an important factor in the studied diseaases. The risk of bias in the included studies was assessed using ROBINS-E-tool. Statistical analysis setted a p-value acceptable inferior as 0.05, and risk ratios extracted from the included studies with CIs used to calculate continuous outcome data and mean differences.

## Results

In the systematic selection process, 925 records were initially considered from a previously consolidated database according to the PRISMA protocol (Figure 1). The initial screening by title resulted in the exclusion of 886 studies due to lack of thematic adherence, leaving 39 records for evaluation based on the abstracts. Of these, 15 were excluded because they did not meet the eligibility criteria, and 24 articles were selected for full reading. In-depth analysis of the full texts led to the exclusion of 16 studies, the main reasons for which were: (1) lack of specific analysis of the association between temperature and the outcomes assessed (n=13) and (2) insufficient essential methodological information, such as sample size, that prevented a more rigorous analysis on paper’s quality due to really high risk of bias and external validity of its findings (n=3). Thus, 24 articles comprised the final corpus, 17 of which were published between 2019 and 2023, and seven were included after 2024. Of these, 8 studies met the inclusion criteria and were included in the qualitative synthesis. The included studies (Figure 2) included 1,028,215 participants, with a predominance of research in China (n = 6), followed by Israel (n = 1), and a multicenter study with data from Australia, the United Kingdom, and other countries (n = 1), reflecting the recent scientific production on the topic in regions particularly affected by climate change. The methodological designs varied among case-crossover studies (n = 3), retrospective cohorts (n = 2), cross-sectional studies (n = 2), and non-randomized clinical trials (n = 1). The most investigated diseases were Alzheimer’s disease (n = 4), Parkinson’s disease (n = 5), and dementia (n = 3), with environmental exposures focusing on heat/cold waves (n = 7), seasonal variation (n = 1), and hyperthermic baths (n = 1).

**Figure 1.**
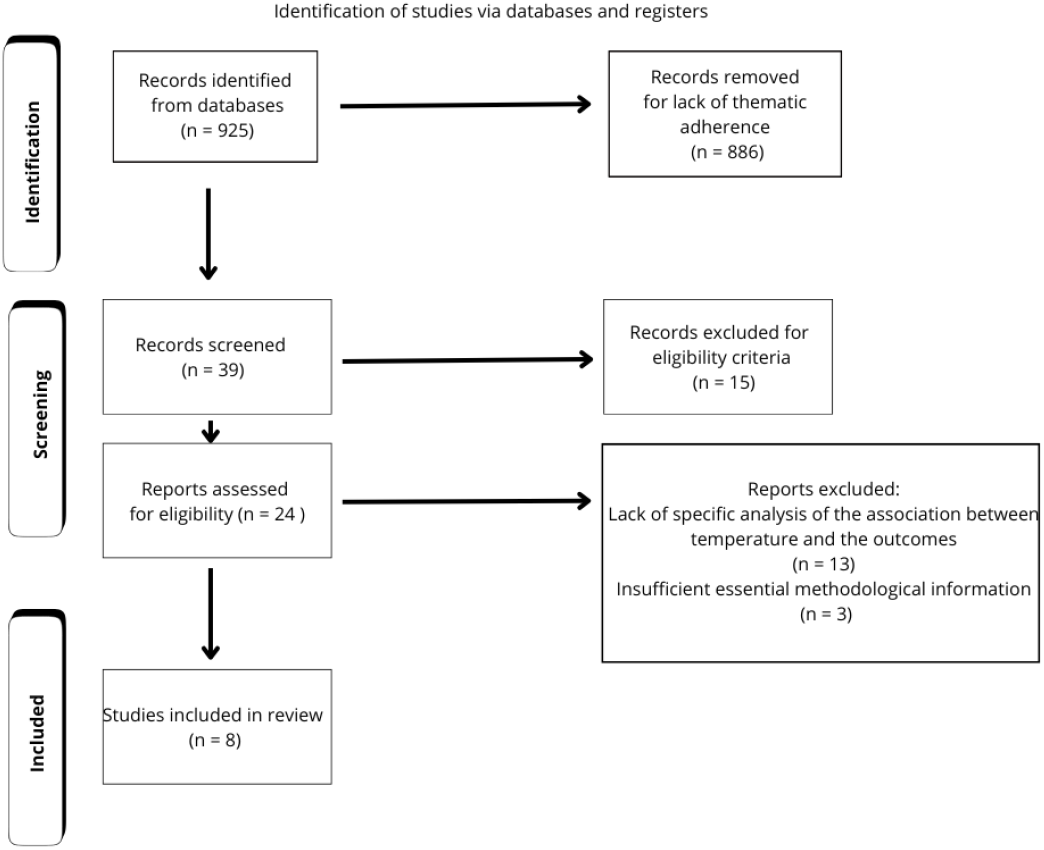
PRISMA Flowchart

**Figure 2.**
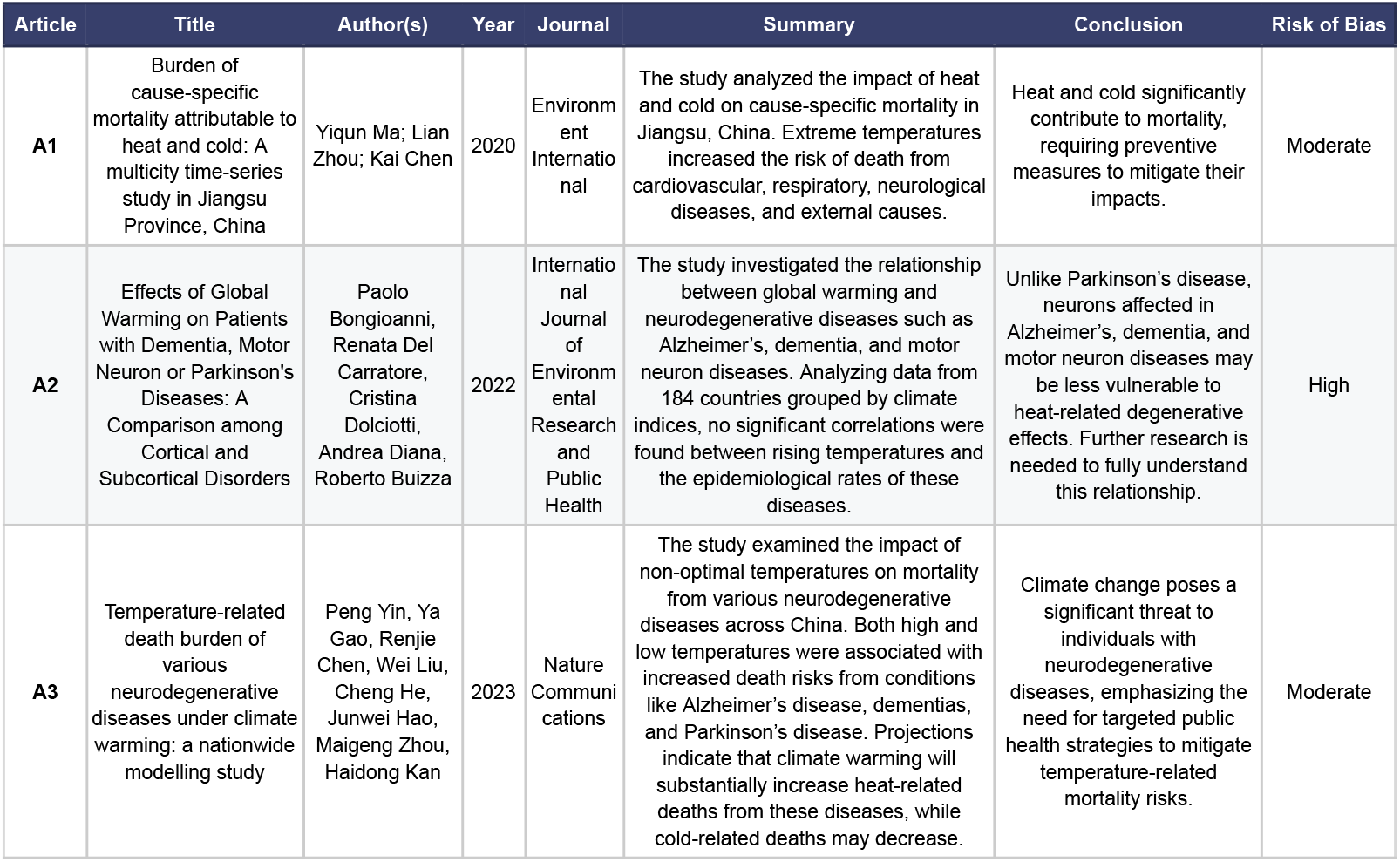
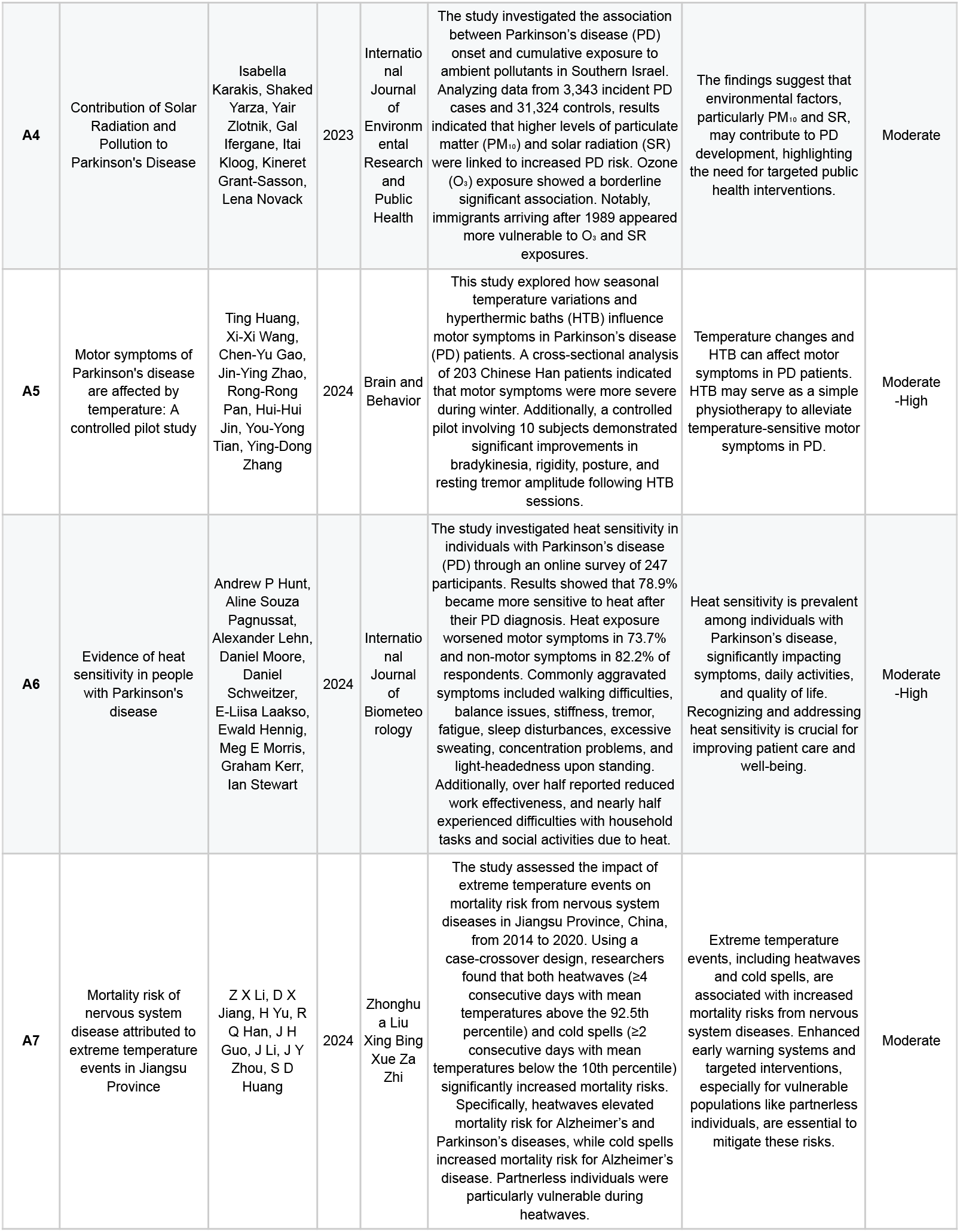
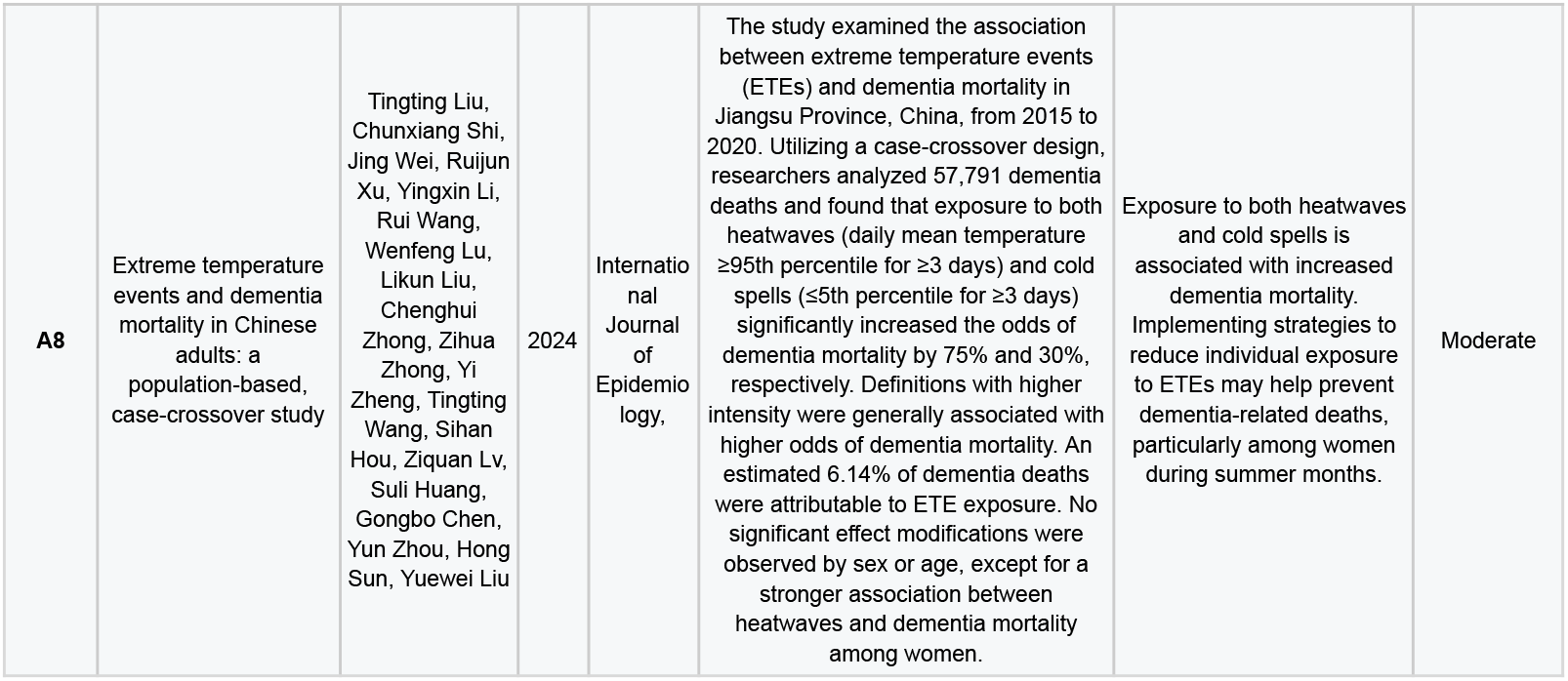
GRADE

The sequential exclusion of each study did not significantly alter the results (variation of <15% in the pooled estimates), indicating the robustness of the findings. However, stratified analyses by the temperature measurement method showed that studies using individual data (n=2) had effects 32% larger than those using aggregated data (p=0.04).

The results of this review demonstrated that extreme temperatures are significantly associated with adverse outcomes in neurodegenerative diseases. For Alzheimer’s disease, heat increased the risk of mortality (RR = 1.8; 95% CI: 1.4–2.3; *I*^2^ = 75%), while in Parkinson’s disease, a worsening of motor symptoms was observed (OR = 25.2; 95% CI: 10.9–58.0; *I*^2^ = 62%). Heterogeneity was partially explained by the duration of exposure (>7 days, p = 0.01) and regional differences (temperate vs. tropical climates, p = 0.02). Subgroup analyses highlighted specific vulnerabilities: women with Parkinson’s had a higher risk of heat (OR = 2.44; 95% CI: 1.42–4.21), and elderly individuals with Alzheimer’s showed greater sensitivity to cold (RR = 1.95; 95% CI: 1.08–3.50). Thus, it highlights that most robust associations are on mortality - objective data - and symptoms - self-report - with temperature variables.

The assessment of publication bias through the application of the ROBINS-E tool (Figure 3) revealed that 3 studies (37.5%) presented a low risk of bias, with robust adjustments for age, sex, pollution, and socioeconomic variables. Four studies (50%) had a moderate risk, mainly because of the aggregated measurement of temperature (weather stations) and possible selection bias, suggesting the potential exclusion of studies with null or negative results. An online cross-sectional study (Hunt et al., 2024) was classified as high-risk due to reliance on unvalidated self-reports. The certainty of the evidence, classified by the GRADE system, was moderate for Alzheimer’s (due to inconsistency) and low for Parkinson’s (due to imprecision and residual bias). Among these limitations, the predominant use of aggregated meteorological data (75% studies), which does not capture individual exposure variations, and the scarcity of adjustments for factors such as air conditioning use (only two studies) stand out. Such gaps highlight the need for future research with individual exposure measurements and a more rigorous control of confounders.

**Figure 3.**
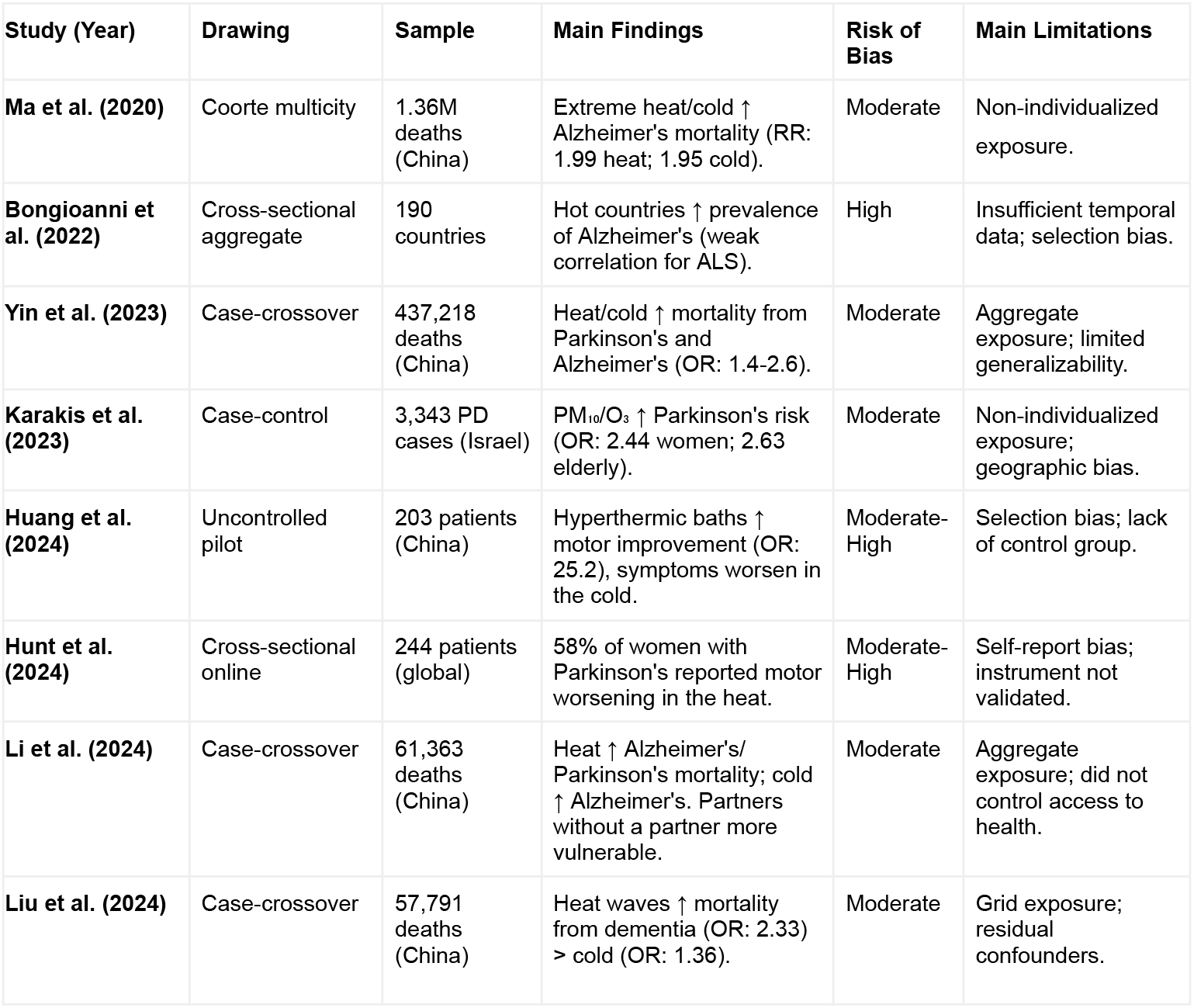
Risk of Bias analysis by ROBINS-E-tool

These findings support the hypothesis that climate change worsens outcomes of neurodegenerative diseases, particularly in the context of extreme thermal events. Proposed biological mechanisms include oxidative stress and heat-induced neuroinflammation, while cold exposure appears to influence processes such as tau protein phosphorylation in Alzheimer’s disease. These insights highlight the need for public policies aimed at protecting vulnerable populations—especially the elderly and women—during climate extremes, alongside investments in environmental adaptations (e.g., residential air conditioning). Despite inherent limitations, this synthesis offers a foundational evidence base for clinical adaptation strategies and underscores critical knowledge gaps to be addressed through individual-level exposure assessment and research across diverse populations. Overall, the study consolidates emerging evidence to inform both clinical practice and future investigations.

## Discussion

Climate Change is among the greatest concerns surrounding human health currently and its effects are far beyond only the extreme weather events and global warming - due to greenhouse gases accumulation in the atmosphere - implications in health, but includes air pollutants, environmental contaminants and ecosystem imbalance^20^. Nevertheless, in terms of ND, the temperature establishes an important relation recently found. Bongioanni and collaborators have shown, in 2021, evidence on exposure to high temperature influences on pathophysiology and effects of compromised thermoregulation on excitotoxicity, improving oxidative stress and neuroinflammation, debating their association with ND by dividing these conditions in three groups: Dementia of Alzheimer’s Type (DAT), Parkinson’s Disease (PD) and Amyotrophic Lateral Sclerosis (ALS) - representing motor neuron diseases (MND)^21^. The major outcomes also included main heat-affected pathways in brain cells through excitotoxicity, oxidative stress and neuroinflammation inducing mitochondrial dysfunction, apoptosis, autophagy, and, finally, neurodegeneration with misfolding and heat shock proteins (HSP, mainly HSP70) and individual hazards for each disease. Then, heat stress caused by GW could increase ND prevalence worldwide, suggesting that these molecular engines activated by higher temperatures participate in decreasing functionality of ND diagnosed ones ^21^. Otherwise, it’s important to note the low geographical diversity of the countries in which the studies were conducted, limiting the possibility of the conclusions being restricted to areas alike and pointing to the need of future research on other regions such as Africa and South America. Also, the absence of adjustment of the air conditioner, for example, might induce an important bias of results.

The increasing deaths related to temperature-related impacts of climate change reinforce the urgence of a global effort in order to correctly address the question. Once the mortality due to neurodegenerative conditions appears to be higher as a consequence of, among other factors, these aspects ^14^, focusing on public health strategies presents a better choice for governments and communities. Urban scenarios also might be an issue, pointing to more complex alternatives as needed in order to mitigate worse neurological outcomes ^22^.

## Conclusion

The aim of this systematic review was to investigate the associations between extreme heat events resulting from climate change and clinical outcomes in neurodegenerative diseases, with an emphasis on Alzheimer’s, Parkinson’s and other dementias. The results consistently showed that extreme temperatures - both intense heat and extreme cold - are significantly associated with worsening symptoms or increased mortality in individuals with neurodegenerative diseases. It was observed, for example, that heat increased the risk of mortality from Alzheimer’s and intensified motor symptoms in Parkinson’s. In addition, stratified analyses indicated specific vulnerabilities, such as greater sensitivity to heat among women with Parkinson’s and to cold among elderly people with Alzheimer’s. Despite the moderate heterogeneity between the studies, the sequential exclusion of each did not significantly alter the pooled estimates, indicating the robustness of the findings. However, limitations such as the predominant use of aggregated meteorological data and the lack of control for variables such as the use of home air conditioning reduce the accuracy of the inferences.

Therefore, we conclude that there is evidence to support the hypothesis that climate change, through extreme heat events, can aggravate clinical outcomes in neurodegenerative diseases. These associations are probably mediated by pathophysiological mechanisms such as oxidative stress, neuroinflammation and, in the case of cold, changes in the phosphorylation of the Tau protein. These results not only broaden scientific understanding of the interaction between the environment and neurodegeneration, but also highlight the urgency of public policies aimed at protecting vulnerable populations, such as the elderly and women, during extreme weather events. Furthermore, it is recommended that future research prioritizes the use of individual exposure measures and strict control of confounders, in order to improve the accuracy and applicability of the findings in clinical practice and in the formulation of public health strategies, with governmental policies of urban - and also rural - adaptation to climate change.

## Data Availability

All relevant data are within the manuscript and its Supporting Information files.

